# Depression and anxiety before and during the COVID-19 lockdown: a longitudinal cohort study with university students

**DOI:** 10.1101/2021.02.23.21252284

**Authors:** Virgínia da Conceição, Inês Rothes, Ricardo Gusmão, Henrique Barros

## Abstract

**Background:** For young people, just as in the general population, COVID-19 caused many changes in their lives, including an increased risk for mental illness symptoms. We aimed to study the impact of the COVID-19 pandemic in anxiety and depression symptoms in a cohort of university students.

**Methods:** This study is part of broader longitudinal research on university students’ mental health with data of the Portuguese version of The Patient Health Questionnaire (PHQ-9) and the Portuguese version of the Generalised Anxiety Disorder (GAD-7) with evaluations on January, May and October 2019 and June 2020, as well as socio-demographic information.

**Results:** 341 university students (257 females and 84 males) were included, with a mean age of 19.91 (SD=1.58). In June 2020, the mean for perceived wellbeing loss was 60.47% (SD=26.56) and 59.54% (SD=28.95) for mental health loss. The proportion of students with scores equal to or above 15 in the PHQ-9 ranged between 22.6% and 25.5% in 2019 and 37.0% in June 2020. The proportion of GAD-7 scores above cut-off ten ranged between 46.0% and 47.8% in 2019 and 64.5% in 2020. Compared with preceding trends, PHQ-9 scores were 3.11 (CI=2.40-3.83) higher than expected, and GAD-7 scores were 3.56 (CI=2.75-5.37) higher.

**Discussion:** COVID-19 impacted negatively depressive and anxiety symptoms, confirming previous studies and young people’s vulnerability in such uncertain times.

## Introduction

In December 2019, the first infection cases with a new coronavirus were reported (Wang et al., 2020a) and soon spread worldwide, becoming a global pandemic. In the first trimester of 2020, most countries initiated a lockdown to prevent the infection from spread.

Although obligatory, quarantine strategies partake in a substantial psychological impact, causing post-traumatic stress symptoms, anxiety, and depression (Brooks et al., 2020). University students present a high incidence of mental disorders (Auerbach et al., 2016), making them a risk group in the current situation.

Concerns about the effects of imposed social distancing on mental health have been growing (Castro-de-Araujo and Machado, 2020; Holmes et al., 2020), and university students’ social interaction and mental health strongly correlate (Elmer et al., 2020). Other significant changes that negatively impact students’ mental health are online teaching, increased worry for themselves and their families, and the future’s economic perspectives (Guessoum et al., 2020). These changes may explain the increased risk of university students’ mental health problems (Solomou and Constantinidou, 2020).

Many studies have researched the mental health impact of the COVID-19 pandemic in the general population (Castelli et al., 2020; Rodríguez-Rey et al., 2020; Thomas, 2020; Wang et al., 2020b). A recent systematic review based on 66 studies estimated that in major depressive disorders, previous prevalence varied from 6.2% to 10.8% worldwide and increased to 31.4% overall due to the pandemic and its quarantine measures (Wu et al., 2021). The same study detected a similar increase in anxiety disorders from a previous prevalence between 10.8% to 14.7% onto an estimated prevalence of 31.9% (Wu et al., 2021).

Most of the studies carried out on the effects of the COVID-19 pandemic analyse the general population, and the studies on university students are still scarce (Marelli et al., 2020; Naser et al., 2020). Another limitation detected in most of the research is the use of cross-sectional data (Gualano et al., 2020; Kaparounaki et al., 2020; Odriozola-González et al., 2020; Rehman et al., 2021), and to the best of our knowledge, only a handful of studies have been carried out with cohort samples, using prospective data that take in consideration mental health problems before the pandemic (Cellini et al., 2020; Elmer et al., 2020; Meda et al., 2020; Pierce et al., 2020).

Cohort studies may minimise sampling bias which can be a particular problem in COVID-19 online sampling (Pierce et al., 2020), and can also be a robust study design to identify better the effects of the COVID-19 lockdown on mental health.

This study’s main aim was to evaluate the impact of COVID-19 lockdown in university students’ anxiety and depression symptoms, taking the opportunity to compare the effects of the lockdown in a cohort previously evaluated with depression and anxiety measures. We also aimed to identify risk factors for developing severe depression and anxiety symptoms in this population.

## Methods

### Participants

In February 2019 (first evaluation moment), all first-year students at the University of Porto were contacted through institutional email and asked to answer a short questionnaire. From those who accepted to participate, 77 were excluded from the sample because their age exceeded the 25-year-old limit marking the end of developmental adolescence characteristics (Sawyer et al., 2018; Twenge and Park, 2019), resulting in a sample of 626 females and 343 males. In May 2019, 415 females and 286 males accepted to be part of a second evaluation moment and, of those, 378 females and 245 males participated in the third evaluation in October 2019. The intervention aimed to reduce depression stigma and improve help-seeking attitudes and took place in May 2019, right before the second evaluation. A description of the sample size in each evaluation moment is available in the supplementary file.

Participants who completed all three 2019 evaluation moments were additionally invited in June 2020 to answer a questionnaire to evaluate the effects of the SARS-CoV-2 pandemic in depressive and anxiety symptoms, resulting in 435 participants.

Depressive and anxiety symptoms were part of the assessment in all four evaluation moments.

### Procedures and outcomes

We asked participants to answer a short socio-demographic questionnaire providing information on sex, age, place of birth, physical health, and previous mental health care. We also asked if students knew anyone that had been infected by COVID-19, with a degree of relationship/closeness, if they had been in prophylactic isolation, satisfaction with social interaction, satisfaction with online teaching, perceived negative impact of the pandemic in wellbeing and perceived adverse effects of the pandemic in mental health.

Participants also answered the Portuguese versions of The Patient Health Questionnaire (PHQ-9) (Ferreira et al., 2018; Sousa et al., 2015) and the Portuguese version of the Generalised Anxiety Disorder (GAD-7) (Sousa et al., 2015). Cronbach’s alpha of the PHQ-9 was 0.86 at baseline, and GAD-7’s Cronbach alpha was 0.91, indicating good internal reliabilities.

We analysed PHQ-9 and GAD-7 scores as continuous variables, indicating a central average score for the sample and a binary threshold score, indicating the proportion of participants with a clinically significant level of symptoms, at least moderate, in need of assessment, and possibly, intervention. As established in the literature, the cut-off point for moderate symptomatology in the PHQ-9 scale is 15 and above (Manea et al., 2012) and 10 in the case of the GAD-7 (Johnson et al., 2019).

Two of the covariates included in the last questionnaire were dichotomic: *“Do you know someone infected with COVID-19?”* and *“Were you on prophylactic isolation?”*. In the case of the first question, we asked participants to identify the individuals they knew with a diagnosis, and the answer options were “parents”, “grandparents”, “siblings”, “other family members” and “friends”.

We also included four more covariates in the last evaluation, asking participants to quantify in percentage, with the aid of visual analogue scales, their satisfaction with social interaction, online teaching, and their perception of COVID-19 pandemic negative impact both on their wellbeing and their mental health. The four questions referred to the last three months and were the following: *“In the past 3 months, how do you evaluate your satisfaction with social interaction?”, “*…, *how do you evaluate your satisfaction with online-teaching”, “*…, *to what extent do you consider that the pandemic has hurt your general wellbeing?”*, and *“*…, *to what extent do you consider that the pandemic has hurt your mental health?”*.

Higher percentages reveal higher levels of satisfaction with social interaction and online teaching in the first two questions. Higher percentages depict the pandemic’s higher negative impact on perceived general wellbeing and higher negative impact on perceived mental health, thus revealing lower perceived wellbeing and lower perceived mental health.

### Data analysis

The statistical analyses consist of two parts. The first part explores the results obtained in the last moment of evaluation, describing depression and anxiety symptomatology after the pandemic and comparing mean scores and above cut-off proportions according to gender and by all the covariates. We used Student’s t-test to compare groups in continuous variables and the Mann-Whitney test to compare proportions between two groups.

The second phase explores the differences in PHQ-9 and GAD-7 scores across time, using One-Way ANOVA repeated measures to assess changes in means and Cochran’s Q test with McNemar’s post-hoc Bonferroni-adjusted alpha to evaluate the changes in cut-off proportions between the different evaluation moments. We used fixed-effects regression to analyse the pandemic’s effect on changes within an individual’s depression and anxiety symptomatology and mean scores for the outcome measure instead of fitting a fixed-effects model for a binary outcome indicator in order to prevent the exclusion with concordant responses over time.

### Ethical and registration considerations

This study is part of comprehensive longitudinal research on first-year university students’ mental health, including an experimental single-blind randomised control trial. (ISRCTN970936), moreover registered as an observational study to analyse the effects of COVID-19 in this cohort (ISRCTN63459073).

It complies with the relevant national and institutional committees’ ethical standards on human experimentation and with the Helsinki Declaration of 1975, as revised in 2008. The Institute of Public Health of the University of Porto ethics committee approved the research with the I.D. reference CE18096. All participants signed an informed consent digital form according to the Helsinki and Oviedo Conventions.

## Results

A total of 341 participants answered all four waves of the study, with a mean age of 19.91 (SD=1.58), ranging from 17 to 25 years old, and 75.4% were female. Compared with the initial sample, the participation rate was 32.06%. However, as we can observe in Table 1, there are no significant differences between participants and dropouts on the main study variables, except for the gender distribution, with a significant decrease in male participation.

**Table 1:**
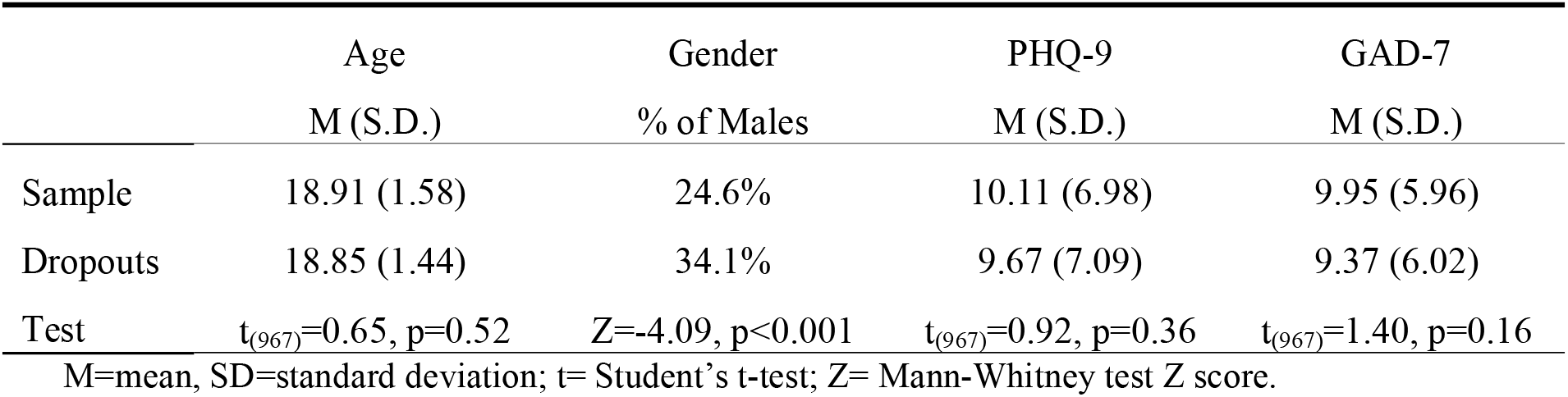
Comparison between sample and dropouts at the moment of the first evaluation.

Most participants did not have to move out of their home residence to go to college (51.6%), 56.6% had never accessed mental health care, and 9.1% lived with a physical illness.

When asked if they knew someone diagnosed with COVID-19, 20.5% (n=70) said yes, and 9.7% (n=33) had to remain in prophylactic isolation. Of those with a close one with a diagnosis, for nine (12.9%) it was a parent, for six (8.6%) a grandparent, for just one (1.4%) a sibling, for 16 (22.9%) other family members, and 38 (54.3%) were friends.

As we can see in Table 2, no differences were identified according to gender, living situation, previous mental health care access or being afflicted by a physical illness in the last evaluation moment.

**Table 2:** Means, standard deviations and above cut-off proportion of PHQ-9 and GAD-7 according to gender, living situation, previous mental health care and physical illness

|  |  | PHQ-9 |  | GAD-7 |  |
| --- | --- | --- | --- | --- | --- |
|  |  | Total score<br>M (S.D.) | Above cut-off (%) | Total score<br>M (S.D.) | Above cut-off (%) |
| Gender | Male | 59.79 (16.17) | 35.7% | 11.35 (SD=6.12) | 61.9% |
|  | Female | 62.68 (18.48) | 37.4% | 12.41 (SD=6.65) | 65.4% |
| | test | $t_{(339)}=0.33, p=0.75$ | $Z=-0.27, p=0.79$ | $t_{(339)}=-1.30, p=0.19$ | $Z=-5.75, p=0.57$ |
| Away<br>from<br>home | Yes | 12.43 (6.69) | 35.8% | 11.72 (6.93) | 62.4% |
|  | No | 13.32 (7.25) | 38.1% | 12.55 (6.50) | 65.5% |
| | test | $t_{(339)}=-1.81, p=0.38$ | $Z=0.44, p=0.66$ | $t_{(339)}=-1.18, p=0.38$ | $Z=-0.78, p=0.44$ |
| Previous<br>mental<br>health<br>care | Yes | 12.76 (6.93) | 35.8% | 12.57 (7.05) | 66.2% |
|  | No | 12.99 (7.05) | 37.8% | 11.82 (6.59) | 63.2% |
| | test | $t_{(339)}=-0.30, p=0.77$ | $Z=-0.44, p=0.66$ | $t_{(339)}=1.04, p=0.30$ | $Z=-0.78, p=0.43$ |
| Physical<br>illness | Yes | 13.71 (7.11) | 45.2% | 12.52 (6.32) | 64.5% |
|  | No | 12.77 (6.96) | 35.9% | 12.08 (6.55) | 64.4% |
| | test | $t_{(339)}=-0.71, p=0.48$ | $Z=-1.02, p=0.21$ | $t_{(339)}=0.35, p=0.72$ | $Z=-0.01, p=0.99$ |

M=mean, SD=standard deviation; t= Student's t-test; Z= Mann-Whitney test Z score.

There were significant differences in satisfaction with online teaching, perception of negative impact on wellbeing and perception of negative impact on mental health between those who knew someone infected and those who did not (Table 3).

**Table 3:** Means and standard deviations of satisfaction with social interaction and online teaching, perception of negative impact on wellbeing and mental health and differences according to ‘knowing someone diagnosed with COVID-19.’

|  | General sample<br>M (S.D.) | Close one infected |  | t-test<sub>(df)</sub> |
| --- | --- | --- | --- | --- |
|  |  | Yes<br>M (S.D.) | No<br>M (S.D.) |  |
| Satisfaction with social interaction | 39.69 (23.72) | 36.86 (29.28) | 40.32 (21.95) | $t_{(339)}=-1.10, p=0.27$ |
| Satisfaction with online teaching | 39.55 (26.60) | 33.23 (26.12) | 41.14 (26.48) | $t_{(339)}=-2.23, p<0.05$ |
| Perception of well-being loss | 60.47 (26.56) | 67.23 (25.13) | 58.73 (26.93) | $t_{(339)}=2.40, p<0.05$ |
| Perception of mental health loss | 59.54 (28.95) | 68.30 (26.87) | 57.28 (29.09) | $t_{(339)}=2.87, p<0.01$ |

M=mean, SD=standard deviation; df=degrees of freedom.

Knowing someone infected with COVID-19 did not affect PHQ-9 or GAD-7 scores. We also did not identify differences between genders in satisfaction with social interaction, with a mean of 36.18 (SD=29.05) in males and 40.73 (SD=21.52) in females (t_(339)_=-1.53, *p*=0.10); a mean of 35.51 (SD=25.28) in males for satisfaction with online teaching and 40.82 (SD=26.89) in females (t_(339)_=-1.59, *p*=0.11); perception of well-being loss with a mean of 55.80 (SD=29.45) in males and 62.00 (SD=25.43) in females (t_(339)_=-1.86, *p*=0.06); nor perception of mental health loss with a mean of 63.51 (SD=31.19) in males and 58.24 (SD=28.13) in females (t_(339)_=-1.45, *p*=0.15).

Comparing the changes in PHQ-9 and GAD-7 scores through time, we observe similar values through February 2019 and October 2019 with a significant increase in June 2020 (Table 2).

ANOVA repeated measures showed significant differences through time: *F*_(3)_ = 14.10, *p* <0.001 in PHQ-9, and *F*_(3)_ = 13.42, *p* <0.001 in GAD-7. Post-hoc analysis showed significant differences only between June 2020 and every other time. No significant differences between PHQ-9 or GAD-7 scores between February and October 2019.

Using 10 as the cut-off points for GAD-7 and 15 for PHQ-9, we verify a significant increase on the above cut-off in the last evaluation (Table 4). Compared with the evaluation on October 2019, in June 2020 we registered more 49 participants with scores above cut-off in PHQ-9 and more 63 in the GAD-7.

**Table 4:** Means and standard deviations of PHQ-9 and GAD-7 scores and cut-off group frequencies.

|  |  | Feb 2019<br>n=341 | May 2019<br>n=341 | Oct 2019<br>n=341 | Jun 2020<br>n=341 |
| --- | --- | --- | --- | --- | --- |
| PHQ-9 | M (SD) | 10.16 (7.01) | 10.53 (7.66) | 9.66 (7.45) | 12.89 (6.99) |
|  | Under 15 | 76.5%<br>(n=261) | 74.5%<br>(n=254) | 77.4%<br>(n=263) | 63.0%<br>(n=214) |
|  | Above 15 | 23.5%<br>(n=80) | 25.5%<br>(n=87) | 22.6%<br>(n=78) | 37.0%<br>(n=127) |
| GAD-7 | M (SD) | 9.94 (5.98) | 9.98 (5.90) | 9.89 (6.19) | 12.15 (6.50) |
|  | Under 10 | 52.8%<br>(n=180) | 52.2%<br>(n=178) | 54.0%<br>(n=184) | 35.5%<br>(n=121) |
|  | Above 10 | 47.2%<br>(n=161) | 47.8%<br>(n=163) | 46.0%<br>(n=157) | 64.5%<br>(n=220) |

M=mean, SD=standard deviation

Cochran’s Q test determined that there was a statistically significant difference in the proportion of students with scores above 10 in GAD-7 (χ^2^(3) = 38.63, *p*<0.001) and above 15 in PH9-9 (χ^2^(3) = 19.48, *p*<0.001).

Post-hoc tests using McNemar’s with Bonferroni-adjusted alpha level of 0.008 (0.05/6) showed that proportions were only significantly different between June 2020 evaluations and any other time for both PHQ-9 and GAD-7. We did not identify other significant differences.

Table 4 shows a higher proportion of moderate to severe cases of anxiety than depression in all evaluation moments. This difference in proportion was significant in all evaluations: McNemar χ^2^ = 67.37, *p*<0.001 in February 2019; χ^2^ = 38.00, *p*<0.001 in May 2019; χ^2^ = 57.79, *p*<0.001 in October 2019 and χ^2^ = 84.79, *p*<0.001 in June 2020.

The fixed-effects regression analysis (Table 5) showed that both in PHQ-9 and GAD-7 last evaluation, all variables had a higher increase than expected, compared with preceding trends. Students with a physical illness increased more the PHQ-9 scores than expected compared to those with no physical conditions. On the other hand, we detected no differences in the anticipated increase within any other model variables.

**Table 5:**
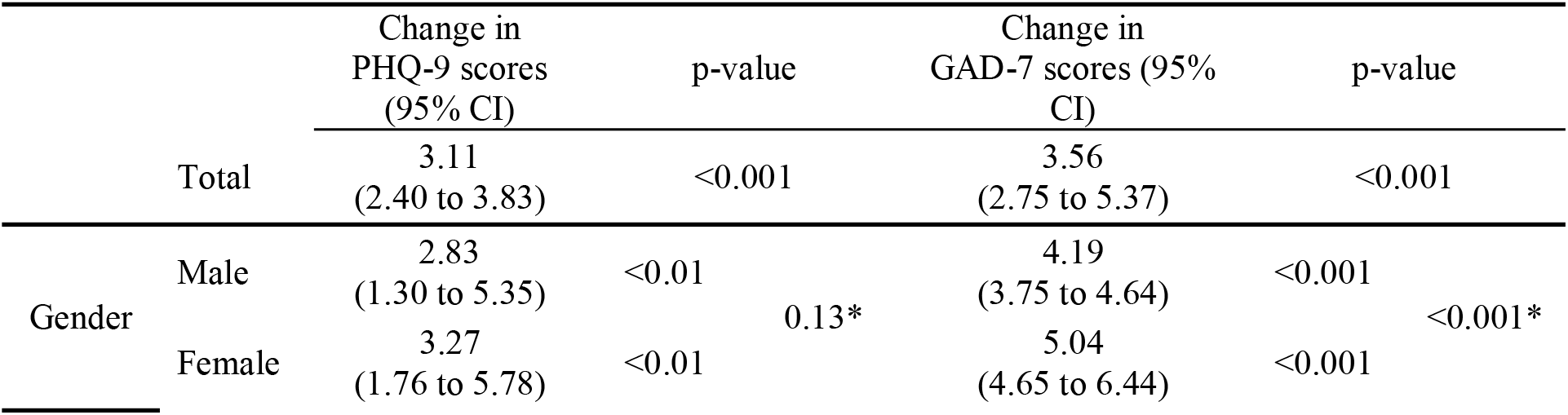
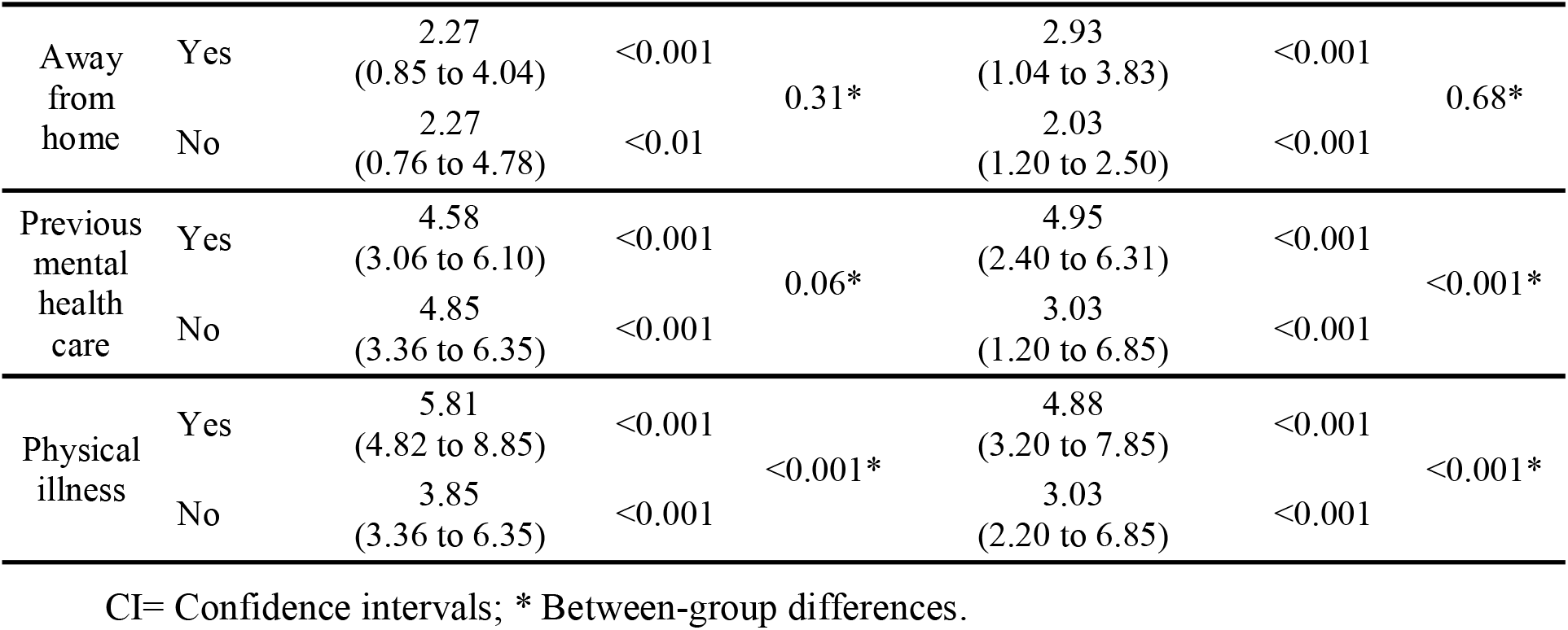
Fixed-effects regression analysis showing the change in PHQ-9 and GAD-7 score associated with the pandemic, compared with preceding trends

In GAD-7, females presented a higher increase mean scores than men; participants who had previously sought help show a higher increase than those who did not, and those with a physical illness show a higher increase than those physically healthy.

## Discussion

The COVID-19 pandemic fostered depressive and anxiety symptoms in university students, thus confirming young people’s vulnerability in such uncertain times (Solomou and Constantinidou, 2020).

Both depression and anxiety symptoms presented stable mean scoring, with no significant changes, between February 2019 and October 2019. However, there was a considerable increase in mean scores and the number of individuals above cut-offs for moderate and severe symptomatology levels in anxiety and depression scales after the COVID-19 pandemic.

About one-fifth of our sample had a close one diagnosed with COVID-19, which may represent a self-selection bias and over-representation of respondents with this experience. The higher percentage of students who knew someone infected may also be due to the more extensive social networking in teenage years (Wrzus et al., 2013). As such, it is expected that young people know more people in general and more so diagnosed with COVID-19, in particular. Most of the known cases were friends, and the definition of friendship is changing with the new online interpersonal dynamics development (Amichai-Hamburger et al., 2013; Nesi et al., 2018), transforming the way young people interact with their peers and widening the range of interpersonal relationships (Nesi et al., 2018) and the number of people considered friends.

Still, knowing someone infected did not seem to affect depressive and anxiety scores, probably because even those that did not know anyone infected lived the fear of contagion and the associated constraints similarly.

Therefore, even if self-selection of more interested people in the COVID-19 and mental health themes occurred, biasing the respondents’ sample, it does not seem to have had a relevant impact since depressive and anxiety symptoms are similar either one knows or not knows someone infected with SARS-Cov2.

Students who knew someone infected also displayed lower satisfaction with online teaching and perceived themselves as more negatively impacted in their wellbeing and mental health. However, as no impact on depression and anxiety symptomatology was observable, that may indicate perceived stress that does not yet translate in scale scores and the possibility of an increase of vulnerability with time, reflecting a need to monitor these young adults.

The number of participants with moderate to severe anxiety symptomatology is higher than the proportion of participants with the same type of depressive symptomatology, which is an expected result since the prevalence of anxiety disorders is more elevated than depressive disorders prevalence (Auerbach et al., 2016). We expected the increase in differences observed between anxiety symptomatology in females and males since females tend to be more vulnerable to mental illness (Solomou and Constantinidou, 2020). Again, many students that have moved closer to the university returned to their homes for the quarantine period, resulting in a more significant change in their day-to-day life. As such, it is not surprising that those students have experienced a higher increase in anxiety levels than those who did not move.

The majority of students have never sought mental health care; however, for those who did so in the past, a higher increase is shown in anxiety scores in the present, which the heightened vulnerability for mental illness after the first episode can explain.

Suffering from a physical illness was the only factor with a higher score than expected for both depressive and anxiety symptoms, compared with their healthy counterparts, identifying the physically ill group as a particularly vulnerable group for mental illness development during the pandemic, which is also not surprising.

These results, specifically the increase of moderate and severe anxiety and depression cases, three months after the first lockdown, confirm the importance of the pandemic negative impact on young adults’ mental health and the need to develop and implement specific prevention strategies targeting and addressing this population needs.

One limitation of this study is the high number of dropouts, increasing the risk of selection bias. The higher dropout rate occurred between the penultimate and the last evaluation moment, corresponding to the final evaluation pre-pandemic and the evaluation post-pandemic, which may be due to increased online activity in general and online research, in particular, developed in these pandemic times. However, there are no differences in the main study variables between participants and dropouts, minimising the risk of bias.

Future research will be essential to explore further the impact on academic performance, social interaction and integration, mental health care service utilisation after the beginning of the pandemic and the type and duration of previous help.

## Data Availability

Data available upon request

