## Supplementary figures and images for "Depression and anxiety before and during the COVID-19 lockdown: a longitudinal cohort study with university students"

### Supplementary file

Figure 1: Sample size description

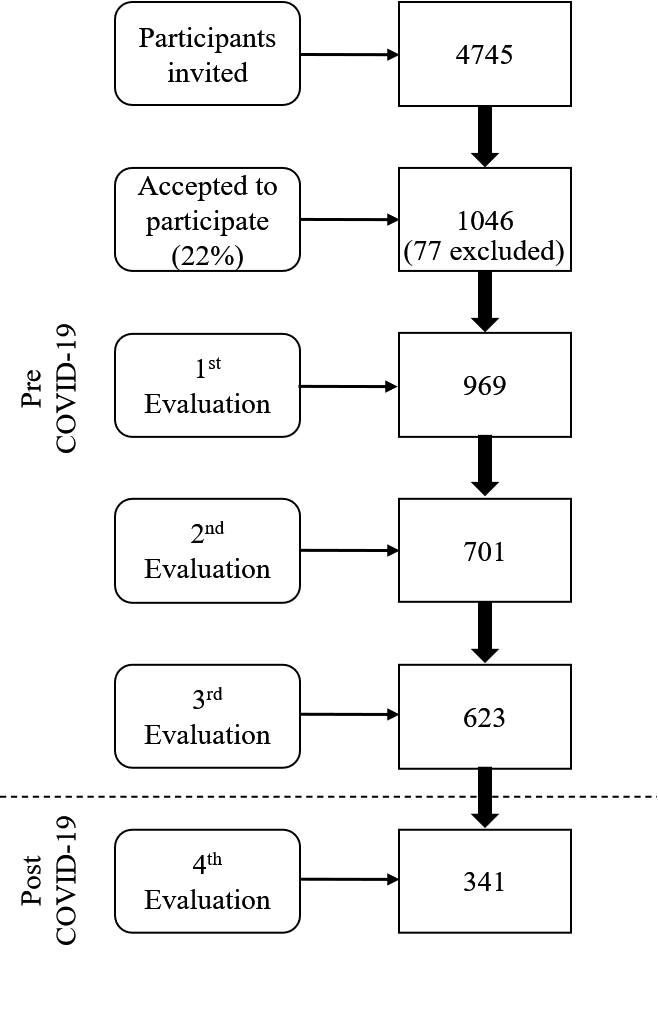
